# Inferring Insulin Secretion Rate From Sparse Patient Glucose and Insulin Measures

**DOI:** 10.1101/2022.03.10.22272234

**Authors:** Rammah M. Abohtyra, Christine L. Chan, David J. Albers, Bruce J. Gluckman

## Abstract

The insulin secretion rate (ISR) contains information that can provide a personal, quantitative understanding of endocrine function. The ISR, if it can be reliably inferred from sparse measurements, could be used both for understanding the source of a problem with glucose regulation system and for monitoring people with diabetes.

**Objective:** This study aims to develop a model-based method for inferring ISR and related physiological information among people with different glycemic conditions in a robust manner.

**Methods:** An algorithm for estimating and validating ISR for different compartmental models with unknown but estimable ISR function and absorption/decay rates describing the dynamics of insulin and C-peptide accumulation was developed. The algorithm was applied to data from 17 subjects with normal glucose regulatory systems and 9 subjects with cystic fibrosis related diabetes (CFRD) in which glucose, insulin and C-peptide were measured in course of oral glucose tolerance tests (OGTT).

**Results:** The model-based algorithm was able to successfully estimate ISR for a diverse set of patients. For CFRD subjects, due to high maximum observed glucose values, we observed better estimation for the degradation rates than for healthy patients. Even though ISR and C-peptide secretion rate (CSR) have different physiological characteristics and nonlinear relations with plasma glucose concentration, our model-based algorithm reproduced the expected linear relationship between ISR and CSR. And the estimated ISR can differentiate normal and CFRD patients: the ISR for individuals with CFRD is substantially lower compared with the ISR for individuals’ normal glucose regulatory systems.

**Significance:** A new estimation method is available to infer the ISR profile, plasma insulin, and C-peptide absorption rates from sparse measurements of insulin, C-peptide, and plasma glucose concentrations.

## 1 INTRODUCTION

Insulin is the essential hormone that regulates cellular energy supply and the intracellular transport of glucose into muscle and adipose tissues (28). The endogenous insulin secretion rate (ISR) quantifies the amount of insulin the body is able to produce as a function of glucose concentration in the blood, providing important information for understanding how an individual’s endocrine system is able to use insulin to regulate glucose regulation. The primary physiological stimulation for insulin secretion from beta-cell is elevated blood glucose levels following nutrition intake and glucose bolus (1). Since direct observations of ISR is not possible, estimating reliable ISR could be useful for diagnosing a problem with the glycemic regulation system and monitoring people with diabetes. The most common methods to estimate ISR utilize plasma insulin and C-peptide concentration measurements.

C-peptide (connecting peptide) is an amino acid polypeptide that is released at the same time, and in the same quantity (their molar ratio of ISR to C-peptide secretion rate (CSR) is 1:1 (13)), as insulin. C-peptide is often used to distinguish insulin produced by the body from injected insulin to estimate ISR, to determine insulin resistance, and to indicate a differential diagnosis of fasting hypoglycemia with hyperinsulinism. C-peptide is released, along with insulin, from the pancreatic beta cells when proinsulin is split into insulin and C-peptide (20, 22). Once insulin and C-peptide are released, they propagate into the liver and kidney. Insulin binds to the liver receptors and starts glucose uptake, inhibiting gluconeogenesis, glycogenolysis, and ketogenesis (3). In contrast to insulin, C-peptide has slight degradation in the liver and is primarily degraded by the kidneys (11). C-peptide also remains in the blood for longer. Whereas insulin is degraded within 15 to 30 minutes (7), while C-peptide takes longer to degrade (14).

Glucose tolerance, insulin resistence, and insulin secretion in a clinical setting are generally measured with various types of glucose tolerance tests. These tests include the intravenous glucose tolerance test (IVGTT) (2), fasting glucose assessment (17, 18), and the oral glucose tolerance test (OGTT) (8). IVGTT are less frequently performed because they are invasive and challenging to endure to the patient and expensive to achieve (16) because of the frequent sampling protocols of the C-peptide up to every minute during an IVGTT. The more commonly used OGTT requires fasting patients to ingest a drink with a fixed amount of glucose and be measured every 15-30 minutes for two to four hours. During OGTT, the plasma and insulin responses reflect the capability of pancreatic beta-cell to produce insulin (19).

Several model-based estimation methods have been developed to estimate ISR. One approach is to estimate from insulin and C-peptide measurements (25, 12, 27, 26). None of these methods assume the ISR is regulated only by glucose concentration but include insulin and C-peptide concentrations. Instead, these multiple compartment methods treat ISR as an unknown time trajectory either without a priori knowledge of its secretion rate function or with different functions to describe the secretion rate. For example, in (12), the deconvolution method is used to estimated ISR by modeling ISR with two exponential functions (biexponential model) with unknown parameters. Another approach that has both one-compartment model (27) and two-compartment model (26) forms is used to estimate the time traces of ISR using a smoothed C-peptide profile generated by cubic spline interpolation. More recently, (25) developed a method to estimate ISR using the Oral C-peptide Minimal Model (OCMM). This method describes the ISR function by two rates proportional linearly with the C-peptide and glucose concentrations. Another recent estimation approach based on OGTT measurements of insulin and C-peptide has been developed to estimate the ISR time traces using two different models, for insulin and C-peptide (21). Our proposed method assumes ISR or CSR depend only on blood glucose (15, 23, 9), and utilizes parametrized fits of these functions in compartmental accumulation models. The fit parameters are optimized to minimize the error between the modeled insulin or C-peptide and measured values.

There has been much effort to use computational modeling of the glucose regulation system to understand normal and disease physiology dynamics, and to infer health of segments of the regulation system from clinical data. Existing models utilize various forms of ISR function with different assumptions and parameters make understanding the glucose dynamics and their relationship to ISR difficult. As illustrated in Figure 1 A, example models have significantly different detailed ISR functions used in these studies (9, 15, 23). In Figure 1 B, will illustrate that these differences yield significantly different glucose responses. A key objective of this work is to provide a methodology to fit this function from clinically recorded data to further improve on this modeling effort. As we see in Figure 1 B, three blood glucose responses are simulated using the model developed by Topp et al. (24) with these ISR functions and subject to the same meal as an input to this model, forming a baseline control. This allows us to understand the function form characteristics, including parameters and shape, change the glucose dynamics significantly in both long-time (B) and short-time (C) courses.

**Figure 1.**
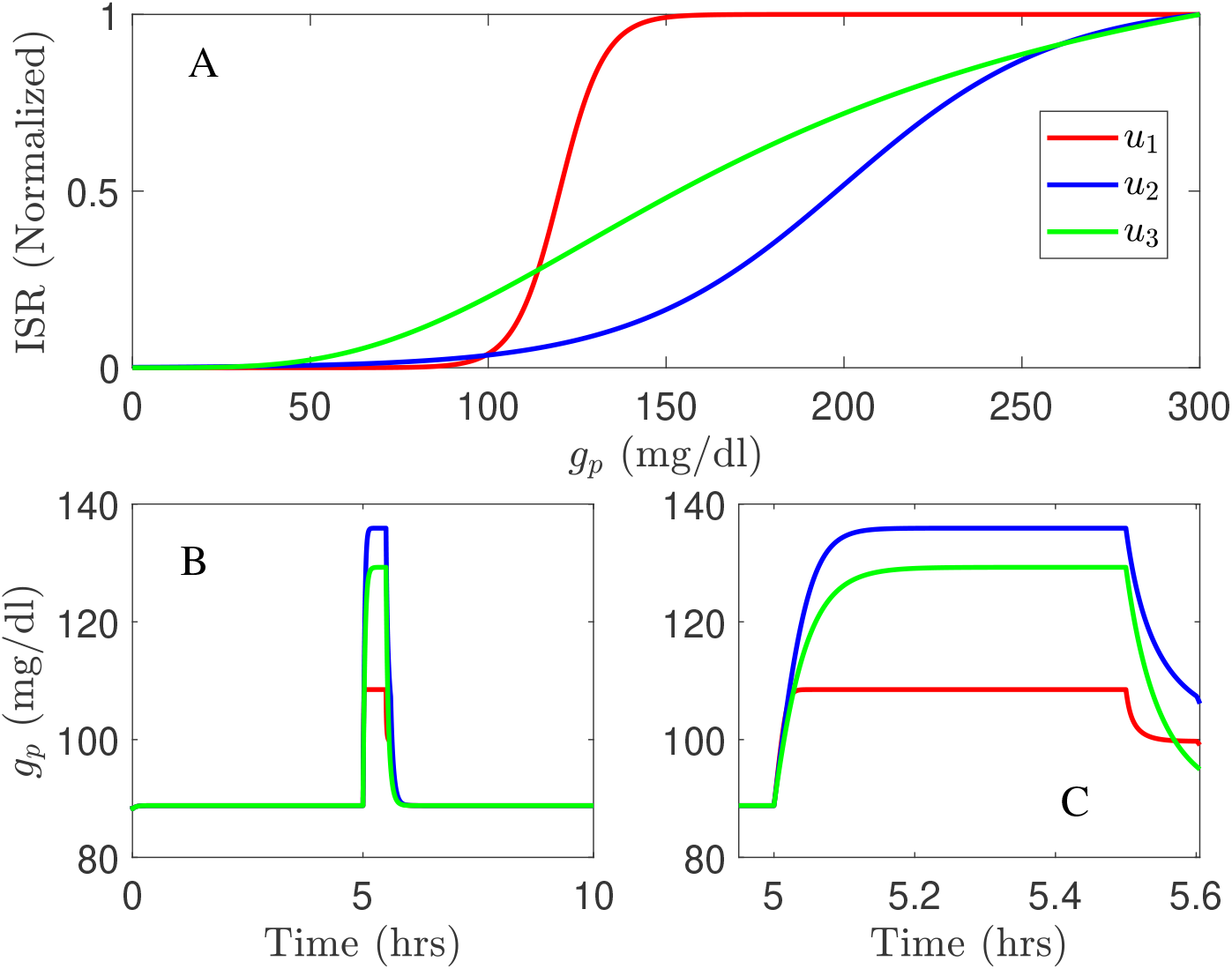
Three different ISR functions (A) generate blood glucose variations, in long time course (B) and short time course (C) simulated using the model developed by Topp et al. (24) with a meal; *u*_1_ is the ISR function adopted from Lui et al. (15); *u*_2_ is the ISR used in Tolic et al. (23), and *u*_3_ is the ISR function used in the model of Joon et al. (9).

To this end, we develop a new estimation algorithm to infer the ISR from OGTT data. Following (15, 23, 9), this new method departs by invoking that ISR is regulated only by blood glucose level. Our approach uses a simple model and independently estimates ISR and CSR from insulin and C-peptide measurements. We use the fact that insulin and C-peptide are produced with a 1:1 molar ratio to validate the output of our estimation results. This result provides physiological insights into beta-cell secretion rates and offers the potential for our approach to treating people with different ISR conditions. We validate the performance of our approach using OGTT clinical data for subjects with different health conditions including, control and CFRD subjects.

## 2 MATERIALS AND METHODS

The proposed algorithm uses parametric models including a single and two compartment models and ISR and CSR function forms with physiological parameters. The parameters of these models and ISR/CSR functions are assumed unknown, but can be inferred from patient data, including plasma glucose, insulin, and C-peptide measurements. We test the performance of this algorithm using OGTT clinical data collected from control and CFRD subjects.

Subjects: Participants ages 6 to 25 years were enrolled as part of GlycEmic Monitoring in cystic fibrosis related diabetes (CFRD) (GEM-CF, NCT02211235), a study of early glucose abnormalities in youth with CFRD. Inclusion criteria for participants with CFRD included a confirmed diagnosis of CFRD by newborn screen, sweat chloride testing, or genetic testing. Exclusion criteria for participants with CFRD included known Type 1 or Type 2 diabetes, use of medications affecting glucose (eg, insulin, systemic steroids) in the prior 3 months, hospitalization in the prior 6 weeks, or pregnancy. For this report, n = 9 youth with CFRD were included. N = 3 (33 %) were male. CFRD individuals were an average age of 14.6 *±* 3.2 years with a mean BMI of 19.0 *±* 2.7 kg/m^2^ and BMI z-score of - 0.28 *±* 0.53. Glucose tolerance categories by OGTT were as follows – 6 CFRD patients had CFRD based on 2h OGTT glucose *>* 200 mg/dL and 3 were classified as NGT. The CF cohort had an average A1C of 5.7 *±* 0.2%.

Healthy controls without CFRD were identified using recruitment flyers and emails at the University of Colorado Anschutz Medical Campus. Exclusion criteria for healthy controls (HCs) included diagnoses of diabetes or prediabetes, overweight (defined as BMI ≥ 85th% by the Centers for Disease Control and Prevention BMI growth charts in youth), chronic disease, acute illness, or pregnancy. A total of n=17 HCs were included of which n=9 (53%) were male. HCs had an average age of 13.3± 3.6 years, BMI of 18.5 ±2.9 kg/m^2^, and BMI z-score of -0.20± 0.68. The HCs had an average A1C of 5.3 ± 0.2%. The study was approved by the Colorado Multiple Institutional Review Board (Aurora, CO), and informed consent and assent obtained.

### 2.1 Insulin and C-peptide Models

The two models, described in Figure 2, are used in the algorithm to reconstruct ISR and CSR. These models, include a single and two-compartment model, which use the same ISR and CSR function but with different parameters, describe the time evolution of plasma insulin and C-peptide. The single compartment model consists of a single plasma pool with a degradation time for plasma insulin and C-peptide. On the other hand,the two-compartment model tracks insulin and C-peptide concentrations in both plasma and interstitial compartments.

**Figure 2.**
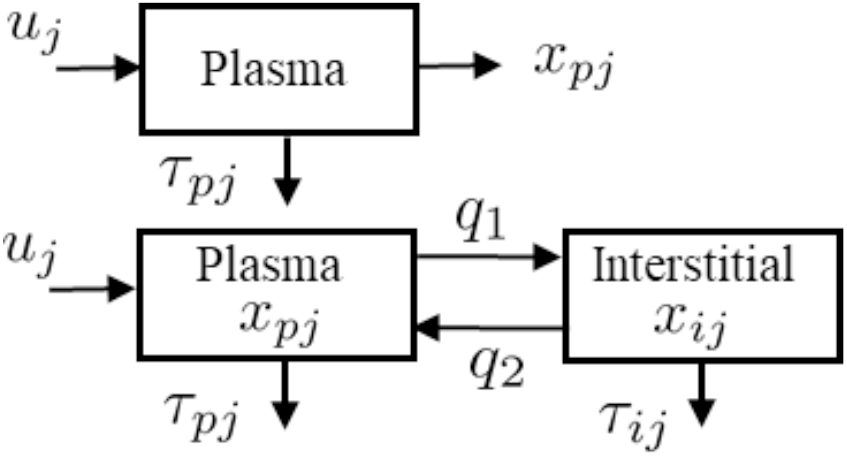
Schematic representations for the single-compartment model (Top) used to describe plasma insulin and C-peptide, and two-compartment model (Bottom) used to describe both plasma and interstitial insulin and C-peptide.

#### 2.1.1 Single Compartment Model

The detail of the single model (Figure 2 Top) is parameterized as follows. The pancreatic beta-cell, which has a nonlinear output secretion function, is denoted by *u*_*j*_, the subscript *j* is an index that takes *I* for insulin and *Cpep* for C-peptide, and releases insulin and C-peptide using various physiological parameters. The subscript *p* denotes plasma, *τ*_*pI*_ and *τ*_*Cpep*_ denote the degradation time for the plasma insulin and C-peptide, respectively. The single compartment model is given by this equation:

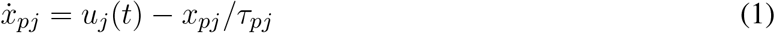

where *x*_*pj*_ is plasma insulin or C-peptide, and *τ*_*pj*_ is the associated degradation time.

Following (23, 15), we use a sigmodal function, which is glucose dependent, for both ISR and CSR, *u*_*j*_, given by

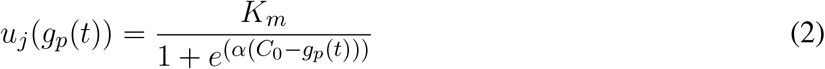

whose parameters (*K*_*m*_, *C*_0_, *α*) are unknown and independently estimated from insulin and C-peptide measurements. In this equation, *g*_*p*_(*t*) (mg/dl) is the plasma glucose concentration at a given time *t* (min), *K*_*m*_ represents a maximum production rate for insulin or C-peptide (*μ*U/ml/min), *C*_0_ refers to a glucose mid-point (mg/dl), and *α* represents 1/width (dl/mg) of the sigmoid curve. We combine the unknown parameters of the single compartment model in this vector Θ_*s*_:

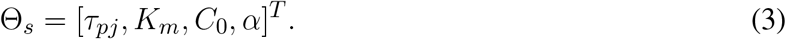

#### 2.1.2 Two Compartment Model

The two compartmental model, as shown in Figure 2 (Bottom), is comprised of two equations:

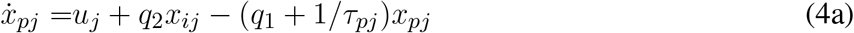

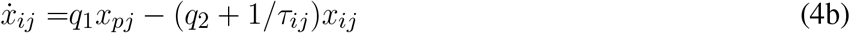

where *x*_*pj*_ and *x*_*ij*_ represent the insulin (or C-peptide) concentrations in the plasma (*p*) and interstitial compartments; *q*_1_ and *q*_2_ represent the mass transport between these two compartments; *τ*_*pj*_ and *τ*_*ij*_ refer to the degradation time for insulin or C-peptide in the plasma and interstitial spaces. The values of *q*_1_ = 0.0473 (min^−1^) and *q*_2_ = 0.0348 (min^−1^) are adopted from the transport model (6). Alternatives to this model include the diffusive transport for example used in the ultradian model (23). We combine the unknown parameters of the two compartment model in this vector Θ_*m*_:

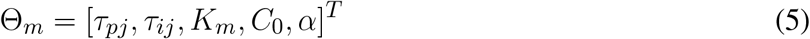

Finally, we provide a summary for the two models given in equations (1) and (4), as follows:

- The accumulation dynamics of the insulin and C-peptide use the same compartment models but with different parameters.
- *u* uses the same function for both ISR and CSR, and this function depends only on the blood glucose values.
- The function of *u* is given by a sigmoid equation (2).
- The parameters of *u*, (*K*_*m*_, *C*_0_, *α*), along with degradation time (*τ*_*pj*_, *τ*_*ij*_), are unknown and estimated independently from insulin and C-peptide measurements.

### 2.2 Compact Form Model

The single compartment model, (2), and the two compartment model, (4), can be formed in a state-space model:

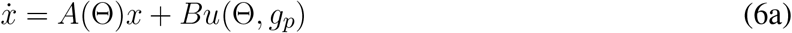

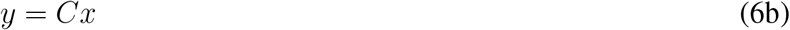

where *x, A, B, C*, and Θ are specific model state and parameters. For the single compartment model (2), we have *x* = *x*_*pj*_, *A* = 1*/τ*_*pj*_, *B* = 1, *C* = 1, and Θ = Θ_*s*_, which is defined in (3). For the two compartment model (4), we have *x* = [*x*_*pj*_, *x*_*ij*_]^*T*^,

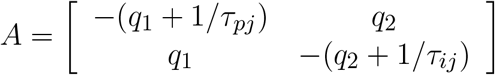

*B* = [1, 0]^*T*^, *C* = [1, 0], and Θ = Θ_*m*_ defined in (5).

Since we use discrete-time data, the state space model, (6), is discretized at a sampling rate of *T*_*s*_ = 0.1min and then given by

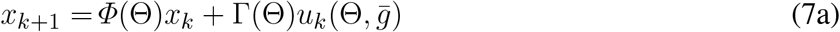

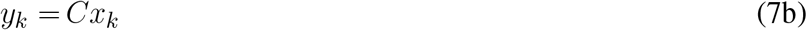

where *Φ* = *e*^*AT*^ and 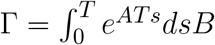. The input *u*_*k*_ is the ISR or CSR, which is a function in both Θ and the interpolated glucose values 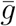 generated from cubic interpolation method.

## 3 RESULTS

The main contribution of this paper is the development of a new estimation approach to infer the ISR from data. The uncertainty in the estimation is studied based on random initial conditions used with the proposed approach to optimize the unknown parameters.

### 3.1 The Estimation Algorithm

Our new estimation method utilizes the above state space model, (7), and is based on the linear least square method (10) to optimize parameters that provide the best fit between the model’s parameters and data. The proposed algorithm uses the interpolated blood glucose values as an input to the algorithm.

The actual time intervals of the measured blood glucose is usually sparse and varies between 10 to 30 min. Given sparse blood glucose measurements, we use the cubic interpolation method to interpolate the blood glucose values between the actual measurements to generate an interpolated glucose trajectory 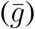 with a shorter time step (*T* = 1 min). This input glucose trajectory is used within the ISR or CSR function 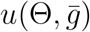 to integrate the model forward generating a model insulin or C-peptide trajectory *y*(Θ, *t*). We take values from this trajectory at *t*_*k*_, which are the exact times as the actual insulin or C-peptide measurements, and use them in the algorithm to optimize the parameters.

For given measurements of blood insulin or C-peptide: *z*(1), *z*(2), …, *z*(*n*), we solve, independently, the following least squares objective function to obtain 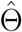:

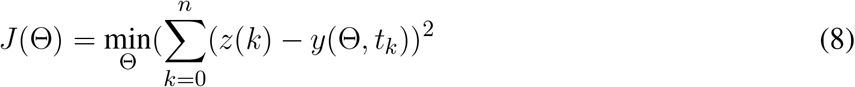

where *y*(Θ, *t*_*k*_) is the model output, which is insulin or C-peptide, generated by 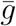, and *z*(*k*) is a measured insulin or C-peptide value. In Figure 3, we provide a schematic representation for our Algorithm 1.

**Figure 3.**
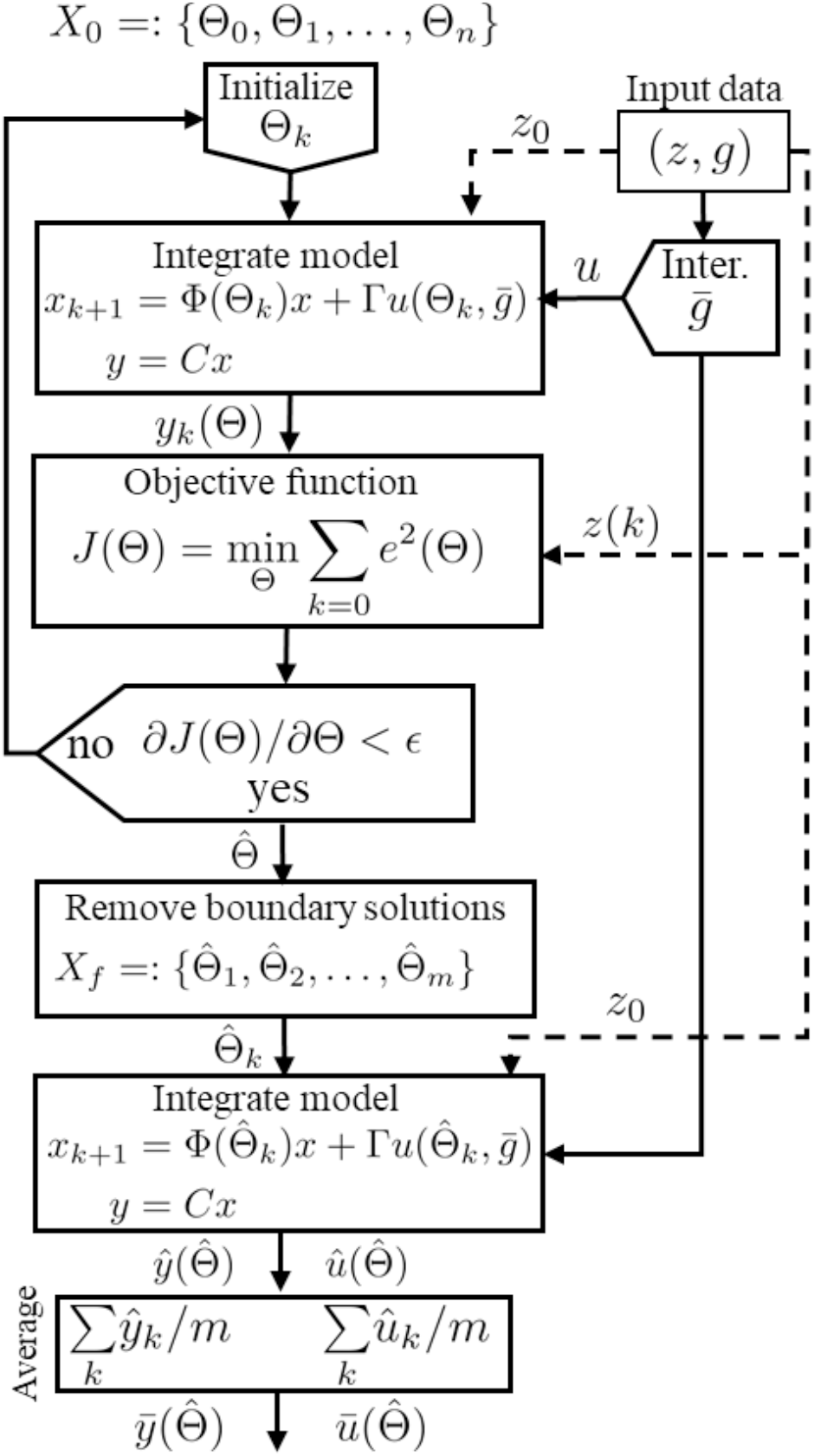
Schematic representation for the estimation algorithm.

#### 3.1.1 Uncertainty Quantification

We explore the uncertainty of the estimated parameter 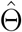 using a randomly sampled initial parameters set *X*_0_ defined with boundaries (lower, upper limits), within plausible ranges. We define these boundaries to exclude all unrealistic solutions and include all possible parameters. In our analysis, for these parameters (*τ*_*pI*_, *C*_0_, *K*_*m*_, *α*), we use the following set of initial parameters defined by *X*_0_ = [10, 180] *×* [200, 1500] *×* [1, 350] *×* [0.015, 0.045]. This set has units as follows, min, mg/l, mU/l/min, l/mg, respectively.

The algorithm’s output generated from this set *X*_0_ is a set of optimal solutions. Not all of these solutions lie on the boundaries of *X*_0_ at the lower and upper limits. We remove all solutions that lie outside the boundaries of *X*_0_ to obtain a refined set of solutions denoted by *X*_*f*_. Each solution in *X*_*f*_ is used within the ISR/CSR function to simulate ISR and CSR trajectories and then generate plasma insulin and C-peptide trajectories by integrating the model, (7), forward using the interpolated glucose values as an input. Finally, we use these trajectories to compute the average and standard deviation (Mean ± SD). These steps are illustrated in Algorithm 1.

##### Algorithm 1

Estimation Algorithm

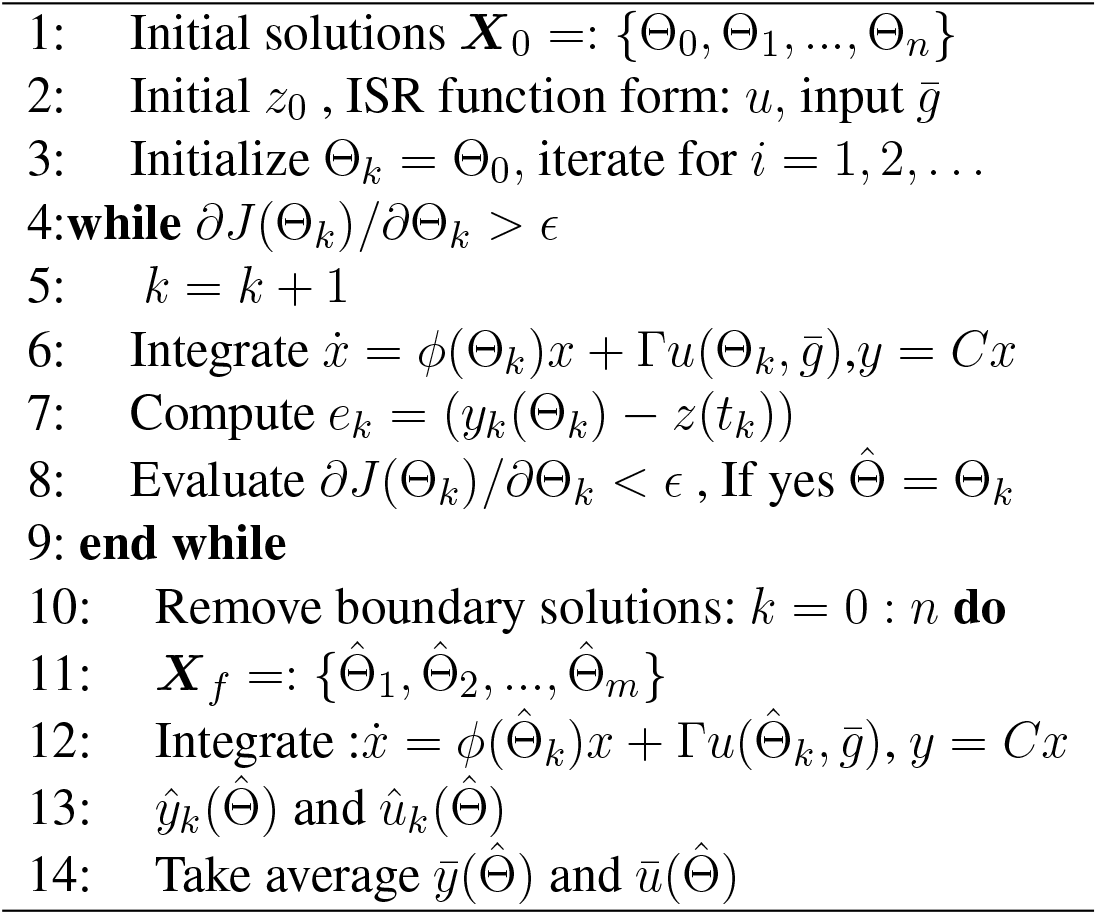

### 3.2 Computational Method Validation

We validate the estimation algorithm using a ground truth generated data from nominal parameters used in (15), with the single compartment model. The nominal values are *K*_*m*_ = 0.6 (mU/l/min), *C*_0_ = 1000 (mg/l), *α* = 0.01 (l/min), and we use multiple values for *τ*_*pI*_ as follows: 10, 15, 30, 60, 90, and 120 min to generate different sets of data from the model. We add 20% noise to each generated data point for both plasma glucose (input) and insulin (output). Each data set is sampled at the same time intervals of the actual data: -10, 0, 10, 20, 30, 60, 90, 120, 150, 180 min, and then we run the algorithm using the initial set *X*_0_ to estimate the nominal values. Therefore, the inferred values of *τ*_*pI*_ matched the ideal (generating) values within an average error of 1.8%, and the other parameters (*K*_*m*_, *C*_0_, *α*) with a roughly average error of 1.2%. This test ensures the performance of the algorithm for estimating ISR parameters and multiple values of *τ*_*pI*_ from noisy generated data.

### 3.3 OGTT Data

We use this method with clinically measured OGTT data, including plasma glucose, insulin, and C-peptide measurements, to estimate ISR/CSR function’s parameters in Θ. These measurements are taken from normal and CFRD subjects at these times: -10, 0, 20, 30, 60, 90, 120, 150, 180 min. Before running the algorithm, the glucose values are interpolated at 1 min intervals using cubic interpolation and then used as an input for estimation. The initial solution *X*_0_ defined in Section 3.1.1 is used with the algorithm and the single compartment model. After estimating the parameter 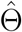 of the single model and following the steps of Algorithm 1, we computed the average solutions over the ensemble, including ISR, CSR, insulin and C-peptide and the histogram for the estimated parameters, for each subject in both control and CFRD groups.

We evaluate the estimation results using the molar ratio between insulin and C-peptide. Insulin and C-peptide are secreted into the blood circulation in a 1:1 molar ratio (13). Therefore, if ISR and CSR are estimated correctly, they should fit a line (e.g., CSR=a(ISR)+b) where a (CSR/ISR) is a slope and b is a constant. To externally evaluate our ISR and CSR estimates, we use the estimated slope between ISR and CSR as a metric to determine how accurate the estimate is. The units for insulin are microunit per milliliter (*μ*U/ml) and for C-peptide measurements are nanograms per milliliter (ng/ml), while the blood glucose is measured in milligrams per deciliter (mg/dl). However, since the insulin and C-peptide measures are not in molar, the slope is *<*1. This slope reflects the (estimated) ratio CSR/ISR conversion factor of (ng/ml-min)/(*μ*U/ml-min), which should be close to the expected value 0.056 of the conversion factor.

In Figure (4), we show estimation results for control and CFRD subjects. The actual glucose measurements (blue circles) and interpolated glucose (magenta), used as an input to the algorithm, are shown in sub-figure A. Also, the measured C-peptide (B, red circles) and estimated C-peptide (B, magenta with standard deviation (± SD) black, green) are presented in the sub-figure B. In addition, the histogram shows the estimated degradation time for both C-peptide (C) and insulin (D). Moreover, the measured insulin (red circles) and estimated insulin (magenta) are shown (with ±SD black, green) in the sub-figure E. Furthermore, the estimated ISR (F, red) and CSR (G, red) are presented with ± SD (balck, green). Finally, the estimated ISR and CSR relationship is shown in sub-figure H. In this figure, the accuracy of the estimation, for both control and CFDR subjects, is indicated by the linear fit between ISR and CSR, as a 1:1 molar ratio. The ISR rates for the control subject (Top, F) are higher than the ISR rates for CFRD subject (Bottom, F). However, the degradation time for insulin and C-peptide for the CFRD subject (Bottom, C, D) is more accurately estimated than the control subject (Top, C, D). We provided physiological interpretation for these results in the next section.

**Figure 4.**
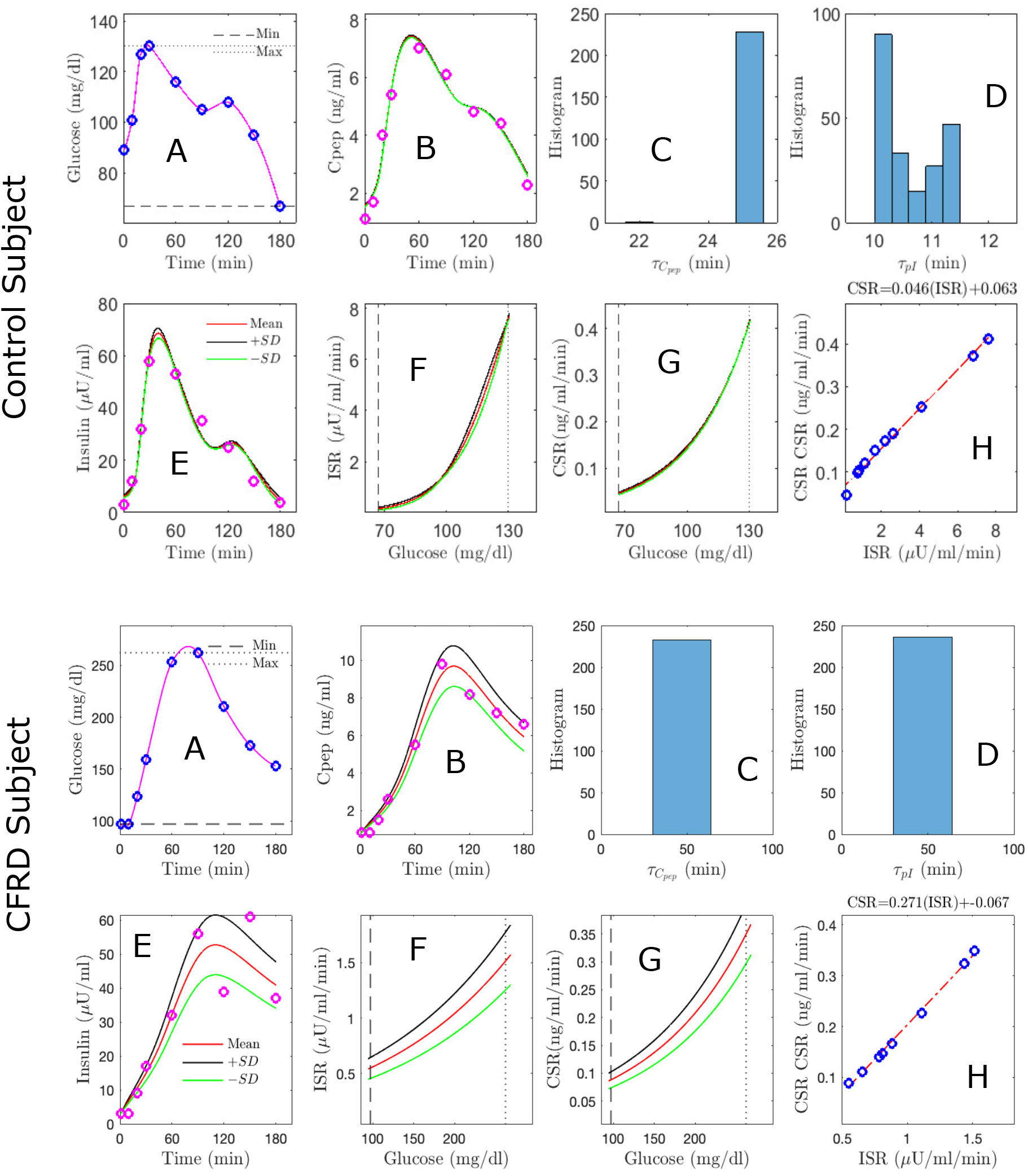
Estimation results, using the algorithm with the single model, for both control (Top) and CFDR (Bottom) subjects, which includes: glucose measurements (A, blue circles) and interpolated glucose (A, magenta) used as an input; measured C-peptide (B, red circles) and estimated C-peptide (B, magenta with standard deviation (± SD) black, green); the histogram is used to represent the estimated degradation time for C-peptide (C) and insulin (D); measured insulin (E, red circles) and estimated insulin (E, magenta with SD black, green); estimated mean ISR (F, red with ± SD black, green); estimated CSR (G, red with ± SD black, green); and CSR/ISR relationship (H, linear fit) indicates the accuracy of the estimation.

Estimation results are obtained and evaluated for all control and CFDR subjects. Our algorithm achieved good estimates of ISR and CSR 13 of 17 control subjects and 7 of 9 CFRD subjects have a good fit as quantified by the observed linear relationship between ISR and CSR and the small mean square error (MSE) between the estimated and observed values of insulin and C-peptide. The patients with poor estimation had a more complex relationship between predicted and observed ISR and CSR. To understand whether these results come from our method or the data, we further investigated these poor estimates.

We diagnose poor estimates by interpreting the blood glucose measurements since our method assumes that glucose only regulates the ISR. In the 4 poorly estimated control subjects, we observed glucose values that had rather severe oscillations starting at 60 min, where the glucose starts from a high value (140 mg/dl) and decreases rapidly to an average level of blood glucose (90 mg/dl) and then fluctuates around 95 mg/dl. This decline in glucose concentration does not allow enough time for the beta-cells to produce insulin, making both ISR and CSR hard to estimate. It is for this reason that estimated ISR and CSR are not well estimated and to not adhere to a linear relationship. In contrast, for in the two poorly estimated CFRD subjects, the glucose value dramatically increased to greater than 250 mg/dl within 40 min. This sharp increase in glucose level reduces the glucose range that produces the whole ISR shape, and beta-cell delivers more ISR in a short amount of time. Consequently, making the the ISR and CSR challenging to estimate correctly.

To test our interpretation for poor estimates, we consider one control subject with an uncertain glucose measurement that dropped at 60 min, from 140 mg/dl to 85 mg/dl. We then removed this 60 min glucose value and the associated insulin and C-peptide measurements. Since we use glucose interpolation as an input to the model, the gap between the glucose values between 30 and 90 min is filled by the interpolated glucose values, as shown in Figure (5) A, B. Without the value at 60min, the insulin and C-peptide measurements are used to re-estimate the parameters. Therefore, we obtain more plausible results for the insulin and C-peptide trajectories (D, F) than the previous estimate (C, E). We also obtain estimation for ISR and CSR values with lower prediction errors. This improvement can be observed by the slope in Figure (H) compared with the previous slope (G). In addition, the MSR between the estimated CSR value and CSR fit (H) is reduced from 0.0287 to 0.0054, and the slope is reduced from 0.061 to 0.054, which is closer to the expected value of 0.056. This suggests that the poor estimate comes from the data, not the method.

**Figure 5.**
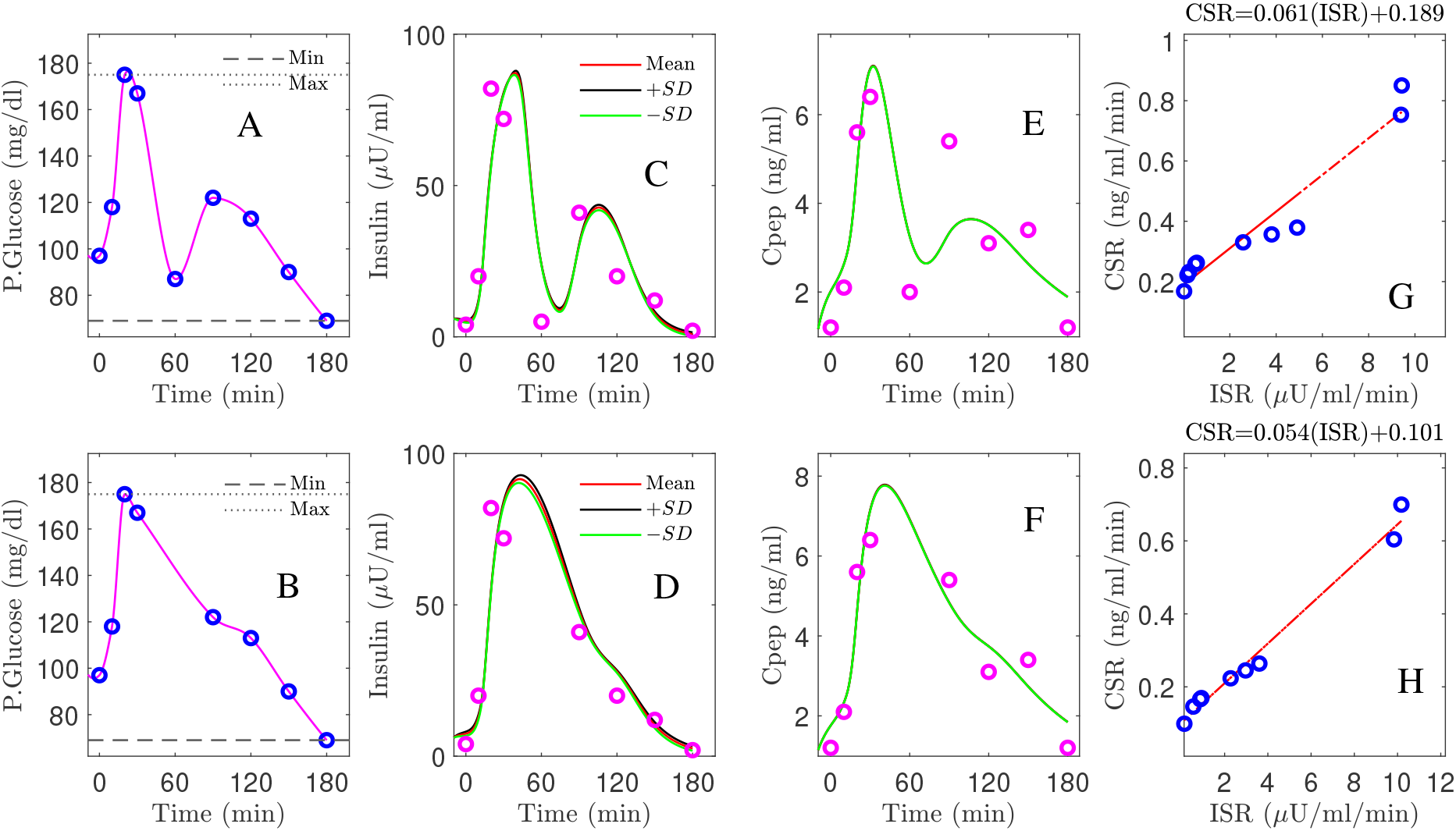
Improvement for the estimation results of plasma insulin (D versus C), C-peptide (F versus E), and slope (H versus G) by removing the uncertain blood glucose value at 60 min (A), and using the interpolated glucose values in the gap between the glucose values at 30, 90 min (B).

In Table 1, we provide a summary for the 13 normal and 7 CFRD subjects who were estimated well, including the ISR average values of the estimated parameters presented by the mean and 95% confident interval, slope, and the ISR evaluated at the glucose value of 140 mg/dl. Note that only 9 subjects of the 13 control subjects have peak glucose values that reached 140 mg/dl, whereas all of the 7 CFRD had blood glucose of 140 mg/dl or greater.

**Table 1.** A summary for the normal and CFRD subjects, including the ISR average values of the estimated parameters, presented by the mean and 95% confident interval, slope, and the ISR evaluated at the glucose value of 140 mg/dl.

| Parameter | $\tau_{pI}$ | $K_m$ | $C_0$ | $\alpha$ | Slope | ISR(140mg/dl) |
| --- | --- | --- | --- | --- | --- | --- |
| Control Subject | 15±7 | 131±61 | 230±45 | 0.06±0.03 | 0.06±0.01 | 150±70 |
| CFDR Subject | 28±12 | 75±60 | 600±240 | 0.014±0.01 | 0.14±0.06 | 73±60 |

We were able to differentiate CFRD from normal patients in two ways. First, as shown in Table 1, the ISR at a glucose of 140 mg/dl is higher for the control subjects than the CFRD subjects. This result indicates the ability of beta-cell for healthy subjects to produce more insulin to mitigate the increased glucose level. Second, the estimated *τ*_*pI*_ for CFRD subjects is larger than the control subjects, as shown in Table 1. This result reflects physiological insights for CFRD that insulin takes longer time to accumulate than in the control subject, to reach a high insulin value, due to a leak in the beta-cells production rate and then takes longer time to drain. Therefore, the increased glucose values in CFRD subjects provide better dynamics for estimation that allows the algorithm to estimate *τ*_*pI*_ more precisely.

The peak of the ISR can be estimated if the glucose range is wide, e.g., 100-350 mg/dl. However, in a short glucose range of 100-140 mg/dl, accurate estimation of the peak is not guaranteed. This observation means it is more likely for CFRD subjects to capture the ISR peak than the control subjects due to the high glucose range in these individuals. We found that two CFRD subjects from among the CFRD group had a glucose range that allowed us to estimate the ISR peak and hence the full ISR functional shape. In these two CFRD subjects, the blood glucose range is between 100-400 mg/dl. On the other hand, the blood glucose range for control subjects is between 95-140 mg/dl, which makes estimating the ISR peak hard to achieve. However, we found only one control subject that the ISR peak was nearly estimated. The glucose range in this control subject is between 95-180 mg/dl. Therefore, we conclude that the peak of the ISR can be better estimated for CFRD subjects, in which the range of blood glucose is wide.

Note that the above fits and figures were obtained using the single-compartment model. However, comparable results can be obtained when incorporating the two-compartment model with the algorithm. But, it is a significant to note that, when the single model cannot estimate the patient’s ISR and CSR, adding a compartment is not helpful. For a comparison between the two models, the ISR was evaluated at the glucose value of 140 mg/dl, and the slope between ISR and CSR was estimated, for control subjects, using our algorithm incorporating the two models. Therefore, shifting to the two-compartment model, the control subjects’ data gives a fraction difference of absolute mean error for the ISR at glucose value of 140 mg/dl provided by (Mean ± SD) is 0.32 ± 0.2. In contrast, the fraction difference of the absolute average error of the slope is given by 0.14 ± 0.15. These results indicate that adding more compartments and unknown parameters is unnecessary to estimate reliable ISR. Instead, a simple model can be incorporated with our method to estimate ISR for people with different beta-cell functions.

## 4 DISCUSSION

We developed a new estimation approach for inferring the ISR from plasma insulin and C-peptide measurements. We validated this method with synthetic data and nominal physiological parameters and were able to reconstruct these values from generated ground truth data with 20% noise is added to each data point for both plasma glucose and insulin. Then the algorithm was applied to OGTT clinical data for both control and CFRD subjects. We use the estimated slope between ISR and CSR to evaluate the estimation for both normal and CFRD groups, as well as the MSE between the observations and the model-estimated values.

We hypothesize that to estimate ISR, it is not necessary to use an OGTT, IVGTT, or other glucose tolerance test. Instead, it can be estimated by knowing glucose values and a nonlinear function of the secretion rate with unknown parameters. Therefore, we specifically implemented a sigmoid function to model the ISR and CSR and then estimate them independently from insulin and C-peptide data. The ISR peak can be estimated, using this function, if the glucose value is higher enough to capture the peak. Using our method, we expect estimate the baseline of the secretion rates if we have more sampled data, especially at the beginning of the test. We use different tests to evaluate the accuracy of the estimation.

Our validation test uses the equal morality ratio between plasma insulin and C-peptide secretion rates. Even though the CFRD and control subjects have different physiology and the ISR and CSR have various physiological parameters and nonlinear relations with plasma glucose concentration, our algorithm recovers the linear relationship between ISR and CSR for both groups. This result indicates the accuracy of the estimation of the algorithm. Another test uses the mean square error (MSE) between the estimation and the measured values. We showed that the estimation, in the two groups, in which the relationship between ISR and CSR is linear, the MSE between modeled insulin or C-peptide and estimated ones is small. This observation reflects the consistency in our results showed by these two validation tests used in our method.

We were able to differentiate the normal and CFRD diabetes phenotypes. We show that the ISR for individuals with CFRD is statistically significantly lower than the ISR for individuals’ normal glucose regulatory systems (see Table 1). However, the ISR peak for the two groups did not differentiate them because the peak ISR was often not observable or computable for normal patients. In addition, due to the high glucose dynamics and slow insulin accumulation in the CFRD subjects, which reveals more information about the insulin degradation time (*τ*_*pI*_), the estimated *τ*_*pI*_ was larger in this group than the control subjects.

To compare the normal and CFRD subjects, we evaluated the estimated ISR at the glucose value of 140 mg/dl for subjects with good estimation in the two groups. After removing the poorly fitted subjects and the control subjects that had not reached the glucose value of 140mg/dl, we obtained 13 control subjects and 7 CFRD subjects out of 17 and 9 subjects, respectively. The results were presented using the empirical cumulative distribution function (ECDF), as shown in Figure (6) A, B. Therefore, we found that the ISR values of the normal subjects were with 50% that the ISR exceeds the rate of 100 *μ*U/ml/min. Whereas, the CFRD subjects were with 50% that the associated ISR value around 15 *μ*U/ml/min. These results indicate that the pancreatic beta-cell of the CFRD cannot produce enough insulin due to the dysfunction of these beta-cells. On the other hand, these beta-cells can produce more insulin at the exact value of glucose (140 mg/dl) in normal subjects.

The estimated slope between the estimated ISR and CSR was also used to characterize these two groups. The slope (CSR/ISR) between the estimated CSR and ISR for both normal and CFRD subjects was plotted in Figure 6 C, D, as ECDF. In both control and CFRD subjects, we observed a straight line describing the physiological relationship between ISR and CSR, with a slope *<*1. The slope is in the unit of ng/*μ*U. When both units are converted to moles, the expected conversion factor is 0.056. In Figure 6 C, D, we plotted this value (0.056) as a vertical (dashed-red) line to illustrate how these slopes, which are predictions of the expected value 0.056, are close to this expected value. As shown in Figure 6 C, the predicted slope of the control subjects was around the expected value of 0.056. On the other hand, the wide glucose range in the CFRD subjects used to estimate both ISR and CSR, which gives more information to estimate ISR and CSR trajectories (e.g., ISR secretion and peak regions), increased the uncertainty in the estimated slope, as shown in Figure 6 D.

**Figure 6.**
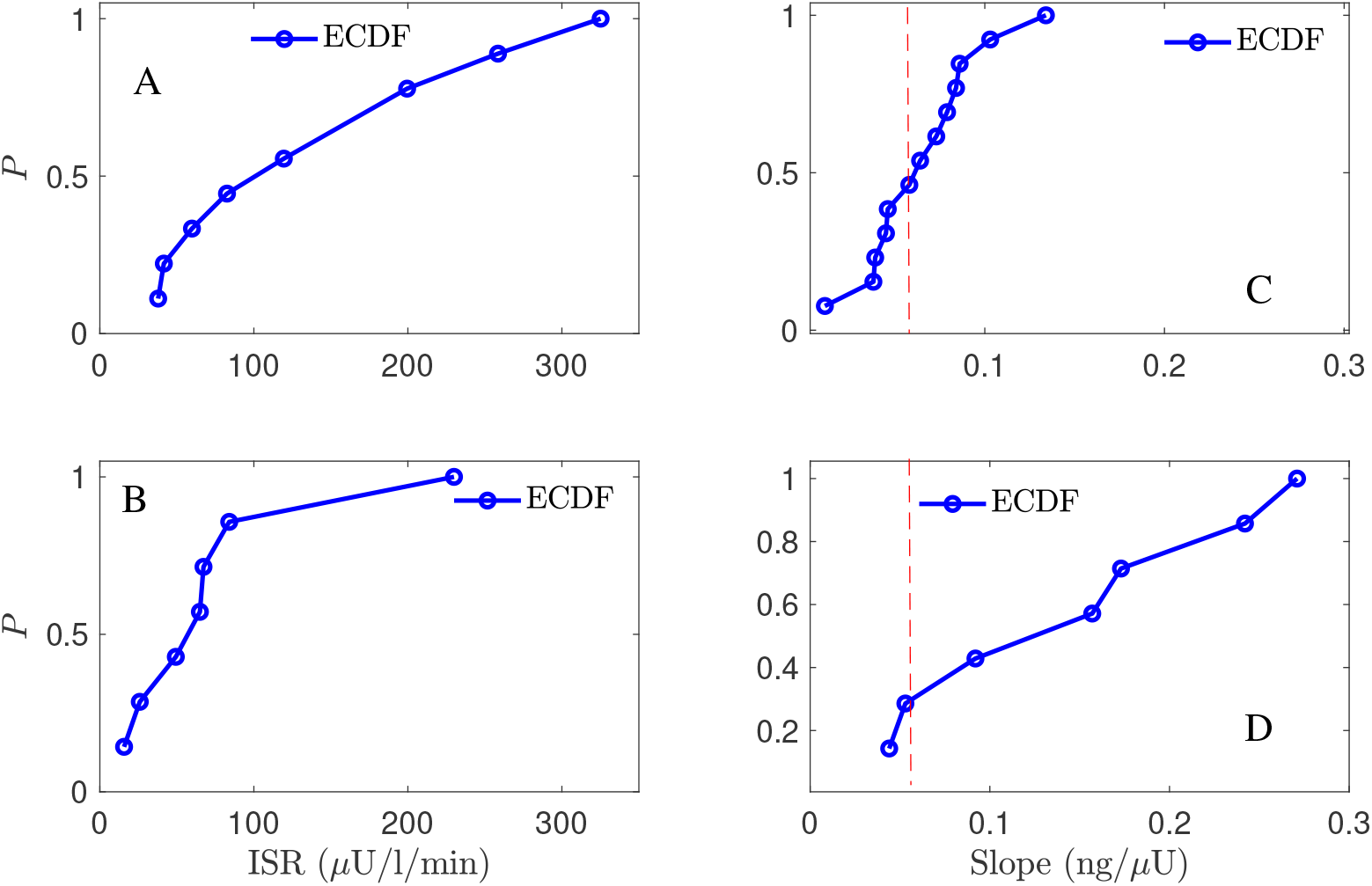
A comparison between the normal and CFRD subjects, using the probability of empirical cumulative distribution function (ECDF), for the ISR value (A, B) evaluated at the glucose value of 140 mg/dl (horizontal) and the slope between ISR and CSR (C, D), the vertical axis is the probability (ECDF); the expected value is 0.056 for the slope (red line C, D).

We may be able to increase testing reliability, the amount of diagnostic information we can extract from a single test, and decrease the need for multiple diagnostic tests using model-based inference. Currently, the ADA recommends for pathways for diagnosing pre-diabetes and type-2 diabetes, one of which includes an OGTT (5). Similarly, the recommendation to diagnose gestational diabetes is via a GTT or OGTT (4). In both cases, a diagnosis requires two tests. By using inference paired with the information in the dynamics of the OGTT rather than a single value, we suspect it would be possible, as we show in this work, to remove inaccurate outliers and accurately estimate ISR and other diagnostic quantities. Additionally, the model provides a platform for extracting additional information. For example, here we estimate the entire ISR curve, increasing accuracy and explainability of the context of the patient state, leading to quantified information regarding how much of the ISR was observed for observed glucose levels and how much excess capacity for insulin production the patient may have, leading to more accurate diagnosis of the patient’s endocrine state.

Considering the above results and discussion, we now have a suitable method with physiological insights about estimating ISR for subjects with different physiological conditions. Furthermore, we showed that using a simple model is good enough to estimate ISR rather than a more complex model with more compartments and unknown parameters. Moreover, we found that using the two compartment, when the single compartment is failed to estimate the ISR and CSR correctly, is not useful. These results allow us to implement the estimated ISR function into glucose models with various fidelity and complicity to understand better the glucose regulation system for patients with different pancreatic beta-cell functions.

## 5 CONCLUSIONS

This study presents a new approach for estimating ISR using plasma insulin and C-peptide measurements. Our approach uses simple insulin and C-peptide models and applies both insulin and C-peptide measures. This algorithm can infer ISR and CSR from OGTT data. Additionally, the method provides a deeper interpretation of the OGTT and a measure of the robustness and accuracy in both the inference and data.

We validate the estimation results in three ways. First, we validate the results by estimating the plasma insulin and C-peptide and comparing MSE between measurements and modeled responses of these variables. Second, we use the 1:1 molar ratio between ISR and CSR to assess the estimation results. We showed thata linear relationship between ISR and CSR can be observed when they are estimated correctly. This result is confirmed in both CFRD and normal subjects. Third, we showed that our algorithm can differentiate between subjects with different beta-cell phenotype-related diseases. Moreover, we showed that the ISR level in CFDR subjects is lower than the ISR level in normal subjects. However, since the variation in blood glucose is high in CFRD patients, the peak of ISR and plasma degradation time of insulin and C-peptide are estimated more precisely. Further, we showed that the estimation of ISR utilizing the single-compartment model is very similar to the results using the two-compartment model. This result indicates the robustness of our approach in estimating the ISR using different models’ complicity and confirms that the ISR can be estimated precisely using only a simple model with less parameters. We also tested our model by treating uncertain measured values in the data. Finally, we provided a physiological interpretation that our method can handle the uncertainty in measured values and improve the estimation of ISR.

The immediate impact of this work is the development of a new approach for estimating ISR, which is now available for determining the beta-cell secretion rates for people with different conditions. This method is ready to implement into glucose models providing a better understanding of the glucose regulation system and monitoring people with diabetes.

## Data Availability

All data produced in the present study are available upon reasonable request to the authors.

